# Neonatal and perinatal mortality in the urban continuum: A geospatial analysis of the household survey, satellite imagery and travel time data in Tanzania

**DOI:** 10.1101/2022.11.14.22282287

**Authors:** Peter M. Macharia, Lenka Beňová, Jessie Pinchoff, Aline Semaan, Andrea B. Pembe, Aliki Christou, Claudia Hanson

## Abstract

**Introduction:** Neonatal mortality might be higher in urban areas. This paper aims to minimize challenges related to misclassification of neonatal deaths and stillbirths, and oversimplification of the variation in urban environments to accurately estimate the direction and strength of the association between urban residence and neonatal/perinatal mortality in Tanzania.

**Methods:** The Tanzania Demographic and Health Survey (DHS) 2015-16 was used to assess birth outcomes for 8,915 pregnancies among 6,156 women of reproductive age, by urban or rural categorization in the DHS and based on satellite imagery. The coordinates of 527 DHS clusters were spatially overlaid with the 2015 Global Human Settlement Layer, showing the degree of urbanisation based on built environment and population density. A three-category urbanicity measure (core urban, semi-urban, and rural) was defined and compared to the binary DHS measure. Travel time to the nearest hospital was modelled using least-cost path algorithm for each cluster. Bivariate and multi-level multivariable logistic regression models were constructed to explore associations between urbanicity and neonatal/perinatal deaths.

**Results:** Both perinatal and neonatal mortality rates were highest in core urban and lowest in rural clusters. Bivariate models showed higher odds of neonatal death (OR=1.85; 95% CI: 1.12, 3.08) and perinatal death (OR=1.60; 95% CI 1.12, 2.30) in core urban compared to rural clusters. In multivariable models, these associations had the same direction and size, but were no longer statistically significant. Travel time to nearest hospital was not associated with neonatal or perinatal mortality.

**Conclusion:** Addressing the higher rates of neonatal and perinatal mortality in densely populated urban areas is critical for Tanzania to meet national and global reduction targets. Urban populations are diverse, and certain neighbourhoods or sub-groups may be disproportionately affected by poor birth outcomes. Research must sample within and across urban areas to differentiate, understand and minimize risks specific to urban settings.

**Key questions:** *What is already known?:*

- Urban advantage in health outcomes has been questioned, both for adult and child mortality
- An analysis of neonatal mortality using Demographic and Health Survey data in Tanzania in 2015-16 showed double risk in urban compared to rural areas
- This phenomenon might be occurring in other sub-Saharan African countries

*What are the new findings?:*

- Categorisation of locations as urban or rural on the 2015-16 Demographic and Health Survey in Tanzania is both simplistic and inaccurate
- Risks of neonatal and perinatal mortality are highest in core, densely populated urban areas in mainland Tanzania, and lowest in rural areas
- Travel time to nearest public hospital was not associated with neonatal or perinatal mortality in mainland Tanzania

*What do the new findings imply?:*

- Extent of urbanicity as an exposure follows a spectrum and needs to be measured and understood as such
- Explanatory models specific to neonatal and perinatal mortality in core urban areas are urgently needed to guide actions toward reducing existing high rate
- Known risk factors such as anaemia and young maternal age continue to play a role in neonatal and perinatal mortality and must be urgently addressed.

## Introduction

Health status and outcomes have generally been described as better in urban compared to rural areas. This urban health advantage is likely due to a variety of factors, including better infrastructure and access to healthcare.^1^ However, this phenomenon is not universal and shows signs of reversal.^2 3^ A recent study of Demographic and Health Surveys (DHS) data collected between 1992 and 2018 in 53 low-and middle-income countries (LMICs) found that the urban advantage in adult mortality has diminished while an urban advantage continues to be observed among children under-five years of age.^4^ The 2020 UN Department of Economic and Social Affairs (UNDESA) World Social Report highlights the extreme inequality that is present within urban areas, as well as relative to rural ones, despite improved resources and infrastructure.^5^

Africa is the most rapidly urbanizing continent, due to natural growth and to a lesser extent urban migration. The continent’s population is expected to double by 2050 and two-thirds of this growth will be in urban areas.^6^ In sub-Saharan Africa (SSA), neonatal mortality has historically been higher in rural areas compared to urban ones.^7^ This urban advantage is likely to be related to a combination of socio-economic factors (maternal education, nutrition, care affordability) and care accessibility (more births occurring in health facilities, shorter distance and travel time to health facilities). With rapid reductions in mortality of children aged 1-5 years, the proportions of under-five deaths have concentrated in infancy, specifically during the neonatal period.^8^ This includes causes of death related to childbirth such as asphyxia, obstructed labour and stillbirths. Recent population surveys have shown that in SSA, the urban advantage in neonatal mortality rate (NMR) might be waning – the most extreme example is Tanzania where urban neonatal mortality (40/1,000 livebirths) is twice the level in rural areas (20/1,000 livebirths). In Tanzania, between the 1990s and 2010s, NMR continued to decline in rural areas whereas it stalled in urban areas. This pattern was also observed in Kenya, Uganda, and Ghana.^7^ Furthermore, the 2015-16 DHS analysis in Tanzania showed that even when available some confounders are adjusted for, and this higher risk of neonatal death persists in urban areas. Within the neonatal period, the disparity between urban and rural is highest in days 1-7 after birth.^7^

The drivers of this observed higher urban neonatal mortality are not well understood; several hypotheses have been proposed and multiple factors could be at play.^7^ To better describe and more accurately explain this pattern, we need to interrogate the concept of urbanicity as the main “exposure”. Much of the existing research frames urban areas as a monolith, whereas there can be significant heterogeneity within. By reducing the urbanicity definition to a dichotomy, studies regarding urban-rural disparities fail to capture urbanicity on a spectrum. Urban areas are not homogenous, and most studies are not able to differentiate between peri-urban and sub-urban areas, areas of informal settlements, urban slums or affluent parts of cities and ignore variations within a single city. A recent study found that 64% of the population in low-income countries reside in small cities or towns, and that more people move to such locations than to larger cities.^9^ In African cities, more than 60% of urban residents live in slums.^10^

Many studies rely on categorizations of urbanicity based on national administrative definitions that are not always an accurate reflection of reality. This is due partly to 1) lack of use of standard criteria, 2) lack of re-evaluation and recategorization of areas over time, and 3) the possible political influence on the categorization (e.g., redefining an area as urban may trigger different requirements regarding government allocation of resources or infrastructure).^9 11^ Some countries use a minimum population size to define urban areas, but the cut-off varies from 2,500 people in Mexico to 20,000 people in Nigeria. However, satellite derived datasets offer an opportunity to use more objective, continuous measures of urbanicity, quantifying the degree of urbanization per grid cell (e.g., 1 km^2^ cells) as a combination of built environment and population density, which is derived independently from national administrative boundaries or designations. Previous studies found that satellite derived urbanicity measures strongly align with administrative data but may fail to capture some rural areas.^12^ By using satellite derived data, multiple categories of ‘urban’ can be derived to validate the observed pattern of higher neonatal mortality in urban areas. It will also allow for an exploration of potential misclassification bias when using country administrative data to define urban or rural compared with satellite derived classification based on population density and land use.

In addition to the misclassification of urban areas, the outcome - neonatal mortality - may also be subject to misclassification with stillbirth due to challenges with establishing whether there were signs of life after birth.^13^ To date, the DHS data does not include full data on stillbirths. Instead, pregnancy history is collected in the contraceptive calendar, and perinatal mortality (i.e., stillbirths and neonatal deaths) can be assessed. However, many key variables on pregnancy and birth, such as place and mode of birth, are not available for pregnancies resulting in stillbirth.^14^ The combination of omission of stillbirths, potential misclassification of birth outcomes (neonatal deaths and stillbirths), misclassification or oversimplification of urbanicity, and residual confounding may mask the true direction and strength of the association between urbanization and neonatal mortality.

With a view to address these limitations affecting our previous work,^7^ we aim to more accurately estimate the direction and strength of the association between urban residence and neonatal mortality in mainland Tanzania using the most recent DHS. We address the limitations by: i) reducing misclassification of exposure by using geospatial techniques to reclassify urban/rural areas, and use a more granular measure of urbanicity (3 categories), ii) reducing misclassification of outcome (neonatal deaths reported as stillbirth) by also examining perinatal mortality (stillbirths and early neonatal deaths combined); and reducing residual confounding by adding several additional variables to the model, including estimated travel time to the nearest hospital.

## Methods

### Overview

We created an alternative urbanicity variable from satellite imagery in lieu of the residence variable provided by the DHS and accounted for the misclassification of neonatal deaths reported as stillbirths. Confounder variables based on literature were retrieved from the DHS dataset and generated through geospatial modelling of travel time to health facilities. We then used bivariate and multi-level multivariable logistic regression models to assess the strength of the association between urban residence and i) neonatal mortality, ii) perinatal mortality.

### Data sources and measures

We used the most recent DHS conducted in Tanzania in 2015-16. DHS are cross-sectional nationally representative household surveys which use standard model questionnaires which countries can adapt. DHS respondents are women of reproductive age (15-49 years), and in several countries men are also interviewed. The surveys include questions on household and individual characteristics, fertility, maternal and child health, mortality, among others. The survey sampling design was based on a two-stage strategy, the first stage involved selection of sampling points (clusters, based on the 2012 Tanzanian census enumeration areas (EAs)) and the second selection of households within clusters. The stratification allowed estimation of certain indicators for 25 regions in mainland Tanzania. Each EA typically contains 20 to 30 households randomly selected to be surveyed from about 100-300 households per cluster. To reduce the disclosure risk, the cluster is first assigned the coordinates of the EA center and then geomasked by displacing the GPS coordinates. Urban clusters were displaced by up to 2 km while rural clusters were displaced by up to 5 km, with a further 1% randomly selected rural clusters displaced by up to 10km.^15^

### Population

Our study population included women aged 15-49 years at the time of the DHS who lived in sampled households and agreed to participate in the survey. We analysed all live births and stillbirths occurring in the five years prior to the survey reported by participating women who had a permanent address in mainland Tanzania.

### Outcome variables

The main outcome of this study was neonatal death. While neonatal deaths are usually defined as deaths between birth and day 28, we also included deaths reported on day 29. This is due to the coding of the response in the DHS dataset and to remain consistent with the cut-off that the DHS report used.^16^ We defined NMR as the number of neonatal deaths per 1,000 live births. We further assessed early (within the first 7 days of life, within which we separated deaths on day of birth) and late (8-29 days inclusive) NMR. The secondary outcome was perinatal death, defined as a combination of stillbirths (defined as deaths of babies at or after seven months of pregnancy and before birth in line with the WHO recommended definition of late gestation stillbirth for international comparisons) and early neonatal deaths. Perinatal mortality rate was expressed as the number of stillbirths and early neonatal deaths per 1,000 pregnancies of gestational age seven or more months, including live births. We extracted stillbirths from the DHS contraceptive calendar based on DHS guidance.^17 18^

### Main exposure

Our primary explanatory variable of interest was *residence* (urban or rural) based on DHS designation and *urbanicity* (core urban, semi-urban and rural) derived from satellite imagery. Globally, there is no uniform definition of an urban area. The DHS relies on the country’s definition of urban/rural which is variable between countries and across time. Statistical offices across countries have used population thresholds of a settlement or a combination of population size and the proportion of residents employed in agriculture to define an urban area.^19^ Specifically in Tanzania, the definition of urban areas is based on all regional and district headquarters and wards with urban characteristics.^20^ Urban wards have above a specified population density and/or a certain percentage of residents in non-agricultural occupations.

As an alternative to the DHS urban and rural classifications, we derived three classes of the urban continuum (urbanicity) - rural, semi-urban, and core urban based on satellite imagery. We used the 2015 Global Human Settlement Layer-settlement model (GHS-SMOD)^21 22^ to classify the location of DHS clusters into different degrees of urbanicity. GHS-SMOD delineates and classifies settlement typologies through cell clusters’ population size, population, and built-up area densities based on the Built-up (GHS-BUILT) areas and Population (GHS-POP) data layers. GHS-BUILT represents the physical extent of the human settlement produced through automatic supervised classification of the Landsat and Sentinel satellite imagery while GHS-POP, a high spatial resolution (250×250 m² and 1×1 km^2^) population density layer is produced by downscaling national census counts data at district level to a regular fine scale grid. It is the combination of GHS-BUILT and GHS-POP based on the *degree of urbanization* concept^23^ that results in GHS-SMOD at the 1 km spatial resolution.^24^

Each pixel of the utilized GHS-SMOD layer^21^ contained a single urbanicity class based on local population density, permanency of the water body, 30% or 50% of a pixel being built-up surface and generalization through smoothing and gap filling.^22^ Based on these rules, level 1 encapsulates 3 classes (urban centre, urban cluster, and rural grid cell) which are further broken down (level 2) into seven classes that were used in this analysis namely; urban centre (class 30), dense urban cluster (class 23), semi-dense urban cluster (class 22), sub-urban or peri-urban (class 21), rural cluster (class 13), low density rural (class 12), very low-density rural (class 11), and water (class 10) grid cells. A detailed description of how these data are generated, processed, and classified is available elsewhere.^22 25^ Supplementary file 1 shows the spatial distribution of the seven classes based on the 2015 GHS-SMOD layer in Tanzania describing the continuum from urban to rural areas in 2015. Urbanicity classes were first reclassified to an ordinal scale from 1 (least urban) to 7 (most urban) after masking out the water class. The average, majority, maximum and minimum values were extracted per buffer and used to define three new classes of urbanicity that were used for the bivariate and multi-level multivariate logistic regression analysis. Buffers were used to reduce the bias associated with scrambling cluster coordinates. We created 2km (urban clusters) and 5km (rural clusters) circular buffers and extracted the polygonal properties of urbanicity as previously implemented^15 26^ and recommended.^27^

Due to the low count of neonatal deaths and homogeneity in some of the extracted urbanicity classes, the seven classes were collapsed into three classes based on the following criteria. Class 1 (core urban) where the mean of all cells per buffer was at least 6, and the maximum and majority of cells per buffer; Class 2 (semi-urban and areas in transition): The mean per buffer is 3 or above but less than 6, or the mean value was less than 3 but had a maximum of at least 4 or above to account for small elements of urban areas such as a small town surrounded by rural areas; the rest of the clusters were assigned to Class 3 (rural areas) that is, where all means were less than or equal to 2 while the maximum values per buffer were at least 4.

### Confounder variables

Potential confounders related to both neonatal/perinatal mortality and urbanicity were identified based on the literature. We relied on confounders available in the DHS capturing the lived environment of the woman (geographic zone), household characteristics, socio-economic characteristics of the woman and variables capturing information about the pregnancy and health-seeking behaviour during index pregnancy and childbirth. Some of the variables were only available for live births and others still only for the *most recent* live birth in the 5-year period (Supplementary file 2).

### Modelling travel time to hospitals

Given that short distances in urban areas can obscure long travel times,^28^ we also included a consideration for accessibility of emergency obstetric healthcare during pregnancy and childbirth generally provided only in hospitals as a potential explanation (effect moderator) between urbanicity and neonatal mortality. A proxy of geographic accessibility to hospital was not available in the DHS and was thus modelled independently for each cluster. It was proxied by the time taken to travel between a DHS cluster and the nearest public hospital, based on a least-cost path algorithm implemented in a Geographic Information System (GIS) via WHO AccessMod 5 software (alpha version 5.7.8)^29^ widely used across healthcare applications in SSA.^30^ We first assembled spatial layers of factors that affect travel time which included ESA Sentinel-2 landcover at 10m x 10m spatial resolution,^31^ road network from OpenStreetMaps (OSM), NASA Shuttle Radar Topography Mission (SRTM) digital elevation model at 30m x 30m spatial resolution, water bodies and protected areas.^32^ The land cover had nine classes (water, flooded vegetation, trees, ice/snow, grass, shrubs, crops, built-up areas and bare ground), while roads were re-classified into four classes (primary, secondary, tertiary, and minor roads) based on OSM description.

The spatial layers were resampled to 300m and merged to form a single layer via the *Accessibility module* in AccessMod software. Travel speeds were then applied on the merged layer to generate cumulative travel time from each cell (pixel) to the nearest hospital in mainland Tanzania at 300 × 300m spatial resolution. Two modes of transport were considered, walking while travelling on off-roads cells, and driving on motorable roads. The adopted modes and travel speeds across the different land cover and road classes were informed by previous studies in similar contexts.^33-35^ Further, walking speeds were corrected for slope derived from DEM using Tobler’s hiking function, an exponential function that describes how human walking speed varies with slope.^36^ The base layer of hospitals was derived from a geolocated pan African master health facility list of public health service providers.^37^ After verification, the list contained 236 public hospitals in Tanzania. The result was a gridded dataset showing travel time to the nearest public hospital in 2015 at 300m spatial resolution. We then linked each DHS cluster with its corresponding travel time to the nearest hospital. Similar to urbanicity, we extracted the average travel time as a continuous variable. For three clusters located in Maisome, Ikuza and Bulyalike islands we used reported travel times from co-author familiar with these regions (ABP). Modelled travel time was not available for these locations because we did not incorporate water as means of transport due to lack of data to parametrise the model.

### Data analysis

We conducted the analysis in three steps. First, we explored the correspondence between the DHS characterisation of clusters as urban or rural in comparison to the three categories based on GHS-SMOD. We also describe the distribution of mean travel time to the nearest public hospital among the study population for both the DHS and GHS-SMOD urban-rural classifications. Second, we described characteristics of the sample and calculated neonatal and perinatal mortality rates, and the distribution of age at death using both DHS and GHS-MOD categorisations. Third, we tested bivariate and multivariable associations between the GHS-MOD urbanicity measure and neonatal/perinatal mortality. The main hypothesis was that there is an association between urbanicity and neonatal/perinatal mortality. Due to inconsistent availability of key variables, we ran four separate multivariable models. The first three models had neonatal mortality as an outcome and were conducted: i. among all live births, ii. among most recent live births, and iii. among most recent live births with newborn’s birth weight and antenatal care (ANC) history available. The fourth model included all births, and the outcome was perinatal mortality.

To assess the effect of urbanicity on neonatal/perinatal mortality, our model building strategy aimed to adjust for confounding, not to overparameterize (i.e., not to introduce unnecessary variables and not to include variables which are on the causal pathway), and to account for any multi-level effects. The selection of variables into adjusted regression models followed previously used approaches.^38-40^ First, based on previous research, we identified all potential confounders (variables that influence both mortality and residence). For each confounder, we ran a bivariate regression to estimate the crude association between each potential confounder and both outcomes (neonatal and perinatal mortality). Only confounders significant at p-value<0.20 were incorporated into the subsequent multivariable regression analysis step.

In the multivariable multi-level logistic regression model, we added urbanicity as the first variable, followed by one confounder at a time, starting with the confounder with the lowest p-value in the bivariate model. Confounders were only retained in the model if they met two criteria; 1) having a p-value<0.05, and 2) effects on the adjusted odds ratio of the confounders already selected (i.e., confounders causing at least a 10% change in the effect size of variables were retained even if not significant at p<0.05). Confounders not meeting these criteria were not retained in the final models except for the geographical zone, which was included *a priori* to capture the lived environment.

Further, we accounted for the multistage sampling design and nesting structure in the DHS data through multi-level hierarchical modelling regardless of the significance.^26 27^ This strategy accounts for contextual factors which are not captured in the fixed variables. We included random intercepts that vary across households and clusters. The household-level random intercept captured the effect of latent household-specific covariates that cause some households to be more similar than others. Cluster-level unobserved characteristics, such as cultural norms, were captured by the cluster random-effect. The choice of cluster and household level random effects was informed by intracommunity correlation coefficient (ICC) tested at the zonal, cluster and household level. Therefore, all four multi-level logistic regression models contained fixed effects and random effects with three levels, clusters at level 1, households at level 2, and individuals (woman-baby dyads) at level 3. We considered variables to be highly correlated if they had a coefficient of over 0.80 based on Pearson correlation coefficient.

Analyses were conducted in Stata Sev15. In all analyses, we adjusted for survey design (*svyset* with clusters, individual sampling weights and stratification). There was no missingness in the urbanicity measure, the main outcomes or other key confounders. There was substantial missingness in the birthweight variable, largely because women reported that their newborns were not weighed.

### Ethics

The DHS received government permission and followed ethical practices including informed consent and assurance of confidentiality. Permission to study this dataset for secondary data analysis was approved by the DHS Program. We did not require separate ethics approval to analyse these secondary datasets.

### Patient and public involvement

Patients and/or the public were not involved in the design, conduct, reporting, or dissemination of this research.

## Results

### Geographic classification of clusters

Mainland Tanzania DHS 2015-16 contained 527 clusters. Based on the GHS-SMOD urbanicity measure, 61 (11.6%) were core urban, 224 (42.5%) semi-urban, and 242 (45.9%) rural. The comparison of DHS and GHS-SMOD classification of clusters is shown in Table 1. All the core urban clusters were correctly identified as urban by DHS. However, there were discrepancies in the other two classes. Among the 224 semi-urban clusters, 138 were reported by DHS as rural and 86 as urban, while among the 242 GHS-SMOD rural clusters, 226 were identified by DHS as rural while 16 were identified as urban. It is expected that the semi-urban classes contain a mixture of urban and rural cells. However, 16 rural clusters were misclassified by the DHS as urban although 13 of these clusters (81%) had majority of the pixels within their buffers as very low-density rural pixels and 9 of these clusters (56%) had maximum values of either 1 or 2. Therefore, these 16 clusters had a very high likelihood of being truly rural.

**Table 1.** DHS Tanzania 2015-16 mainland clusters based on DHS versus GHS-SMOD classification.

|  |  | GHS-SMOD urbanicity classes |  |  | Total |
| --- | --- | --- | --- | --- | --- |
|  |  | Core urban | Semi-urban | Rural |  |
| DHS residence | Rural | 0 | 138 | 226 | 364 |
|  | Urban | 61 | 86 | 16 | 163 |
|  | Total | 61 | 224 | 242 | 527 |

### Travel time to nearest hospital

The average travel time from each cluster to the nearest public hospital was 63 minutes, with large sub-national variations at high spatial resolution. At cluster level, modelled travel time estimates ranged between 0 and 418 minutes (7 hours). Among the 527 included clusters, 349 (66%) were within a 1-hour catchment of the nearest public hospital, while 23% (121 clusters) were within 2-3 hours (Supplementary file 1). Stratification by urbanicity showed that the DHS rural and urban classes had an average of 14 and 78 minutes, respectively. In the three new urbanicity classes, the average travel time was 89 minutes in rural clusters, 41 minutes in semi-urban clusters and 4 minutes in core urban clusters. Majority of urban and core urban clusters were within 30 minutes of the nearest public hospital (Figure 1).

**Figure 1.**
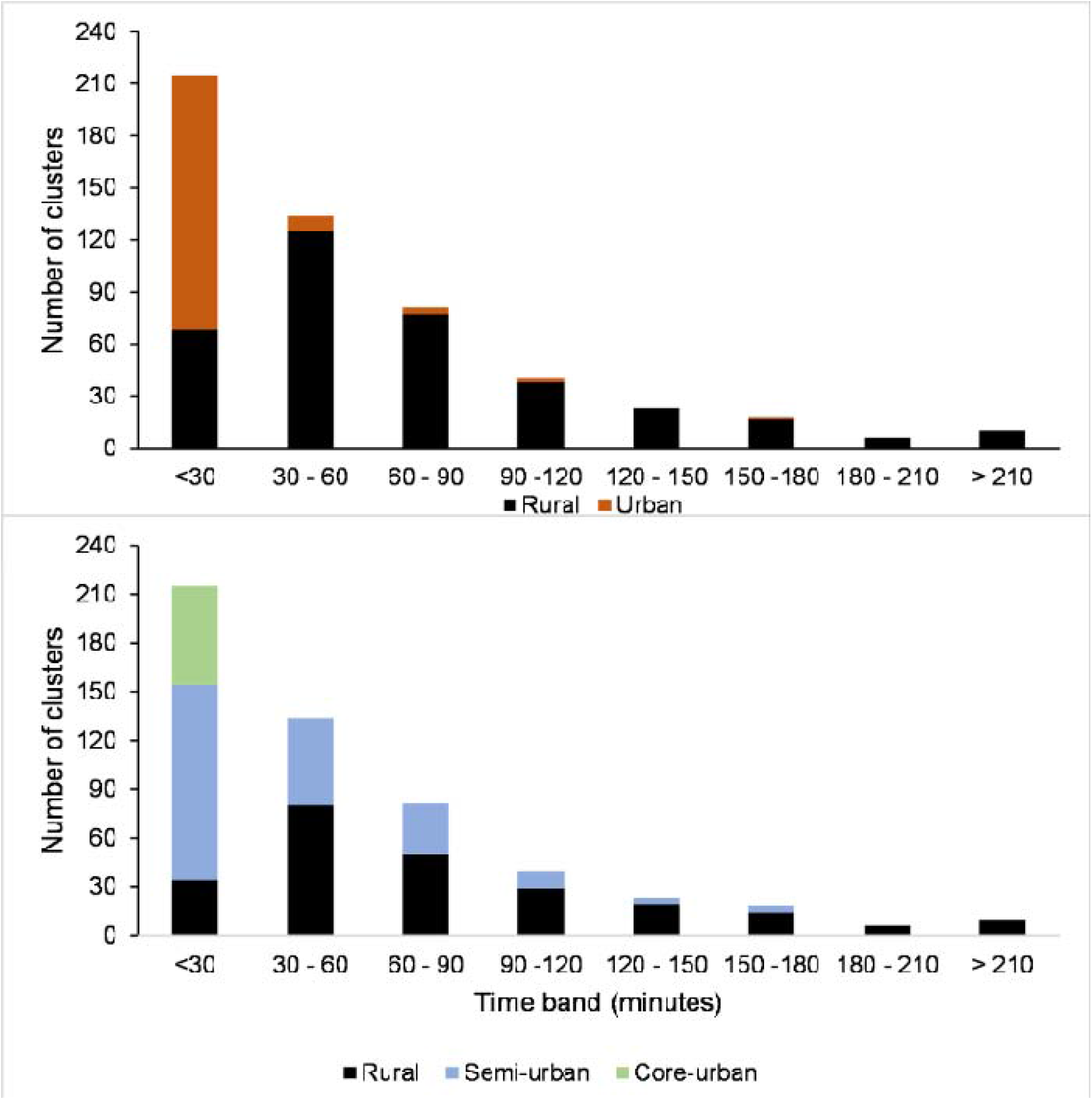
Distribution of 527 clusters in mainland Tanzania by travel time to nearest public hospital in minutes by the DHS (top panel) and GHS-SMOD (bottom panel) urban classification of clusters

### Description of the sample

The analysis dataset contained 8,915 pregnancies of seven or more months among 6,156 unique women: 3,765 women contributed one pregnancy, 2,042 women contributed two pregnancies, 330 contributed three pregnancies and 19 contributed four pregnancies. Among these 8,915 pregnancies, 8,739 resulted in live births and 176 in stillbirths. Among the live births, 217 neonatal deaths were reported (180 early neonatal and 37 late neonatal). A total of 356 perinatal deaths (stillbirths + early neonatal deaths) were reported. Table 2 shows the distribution of the outcome variables and the characteristics of the analysis sub-groups based on availability of variables, for Tanzania mainland overall and by GHS-MOD urbanicity categories. More than half of all births in the sample occurred in core urban or semi-urban areas.

**Table 2.**
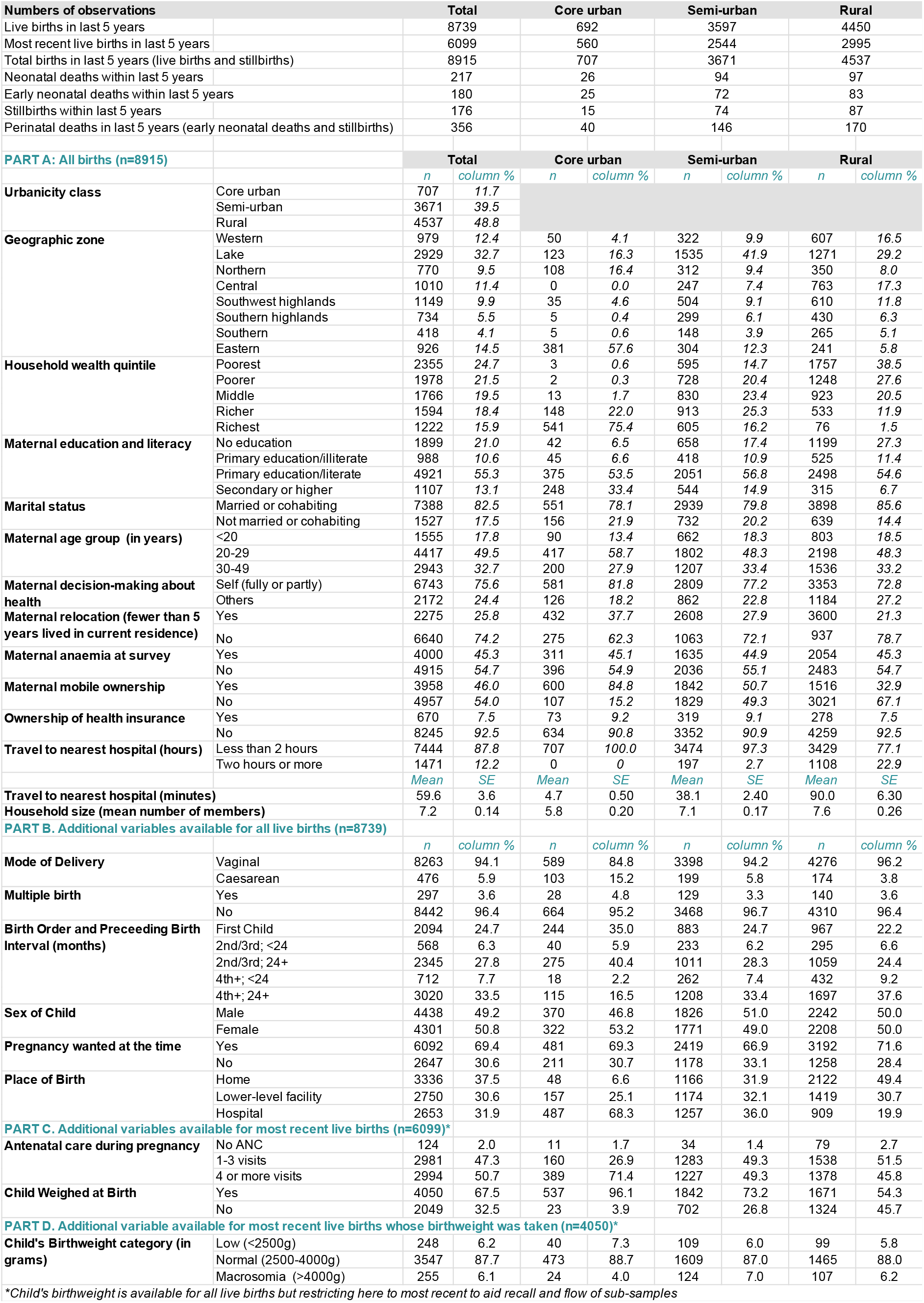
Characteristics of sample of pregnancies and births in analysis, overall and by urbanicity class.

### Neonatal and perinatal mortality

Table 3 presents the neonatal and perinatal mortality rates by DHS residence, GHS-SMOD urbanicity classification, and for mainland Tanzania overall. The comparison shows that mortality estimates for rural areas did not differ between DHS and GHS-SMOD classifications. The perinatal and neonatal mortality rates in the new urbanicity class of semi-urban were similar to levels in rural categories of both DHS and GHS-SMOD. Within the GHS-SMOD classification, core urban areas reached the highest perinatal (56.4/1,000 pregnancies) and neonatal mortality rates (39.8/1,000 live births); these were significantly higher than those observed in semi-urban and rural areas.

**Table 3.**
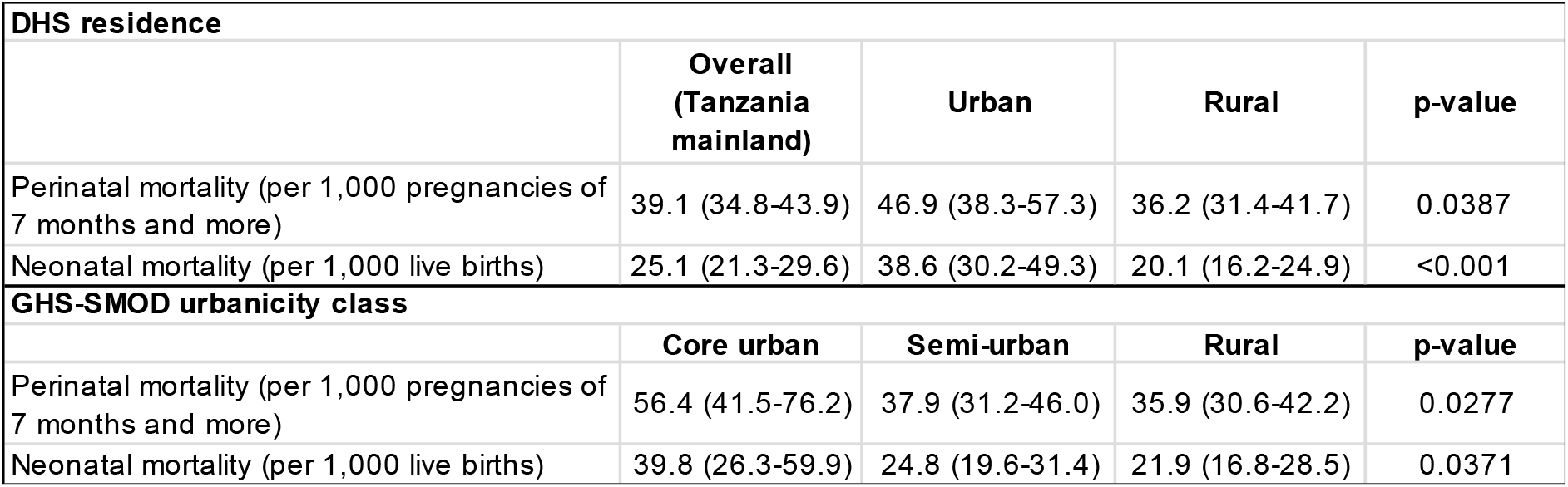
Neonatal and perinatal mortality rates by DHS urban/rural residence and GHS-SMOD urbanicity categories, with 95%CI.

Further details of neonatal and perinatal mortality are shown in Supplementary file 3. Briefly, among the 217 neonatal deaths, the distribution of timing of death was significantly different by urbanicity; in core urban clusters, more than 95% of neonatal deaths occurred in the first week of life (predominantly on days 2-7), compared to 19% in semi-urban and 14% in rural areas. However, within the early neonatal period, semi-urban and rural areas had a higher percentage of deaths on day of birth compared to core urban. The mean age at death was 4.1 days; this was shortest in the core urban category of clusters (2.9 days) compared to semi-urban (5.1) and rural (3.6). Among the 73 most recent neonatal deaths of babies born in facilities, we looked at whether the death occurred before or after discharge from the facility. Two fifths of neonatal deaths in core urban and rural clusters occurred after discharge; this was much higher (73%) in semi-urban areas, a significant difference despite the small sample size; but corresponding with the results on distribution of time of death. We also examined the ratio of stillbirths to early neonatal deaths which is a proxy for misclassification between two outcomes and stillbirth data quality. The ideal ratio should be around 1.2 with much lower or higher values indicating possible underreporting or misclassification.^41^ Overall, among all areas in Tanzania the ratio was 0.85 indicating a small degree of underreporting. However, when examined according to urbanicity status, coreurban areas had the most underreporting or misclassification of stillbirths with a ratio of 0.52 compared with semi-urban and rural areas which had reasonably good ratios just below 1.

### Bivariate analysis

Bivariate analysis examining the association of variables with neonatal death and perinatal death is shown in Table 4. Compared to GHS-SMOD rural class, the odds of death were not higher in semi-urban areas, but were significantly higher in core urban areas (OR=1.85, 95%CI 1.12-3.08) for neonatal death and 1.60 (95%CI 1.2-2.3) for perinatal death. Compared to the Lake zone, only Southern and Eastern zones had significantly different (higher) neonatal and perinatal mortality. Women from richest households had higher odds of reporting neonatal mortality. More educated women (primary education and higher) had higher odds of reporting neonatal mortality compared to women without formal education, but this association was not noted for perinatal deaths. Compared to women 20-29 years of age at time of index birth, women <20 years had 50% higher odds of reporting both neonatal and perinatal mortality. Maternal anaemia at time of survey was associated with 35% higher odds of both neonatal and perinatal death. Maternal decision-making, insurance coverage, mobile ownership, and travel time to nearest hospital were not associated with either outcome. Larger household size was associated with lower odds of both outcomes. Among live births, the odds of neonatal death were higher for caesarean mode of delivery, multiple births, primiparous mothers, male newborns, hospital births, and lack of ANC during pregnancy. Among newborns who were weighed, both low birthweight and macrosomia were associated with higher odds of neonatal mortality compared to normal birthweight.

**Table 4.**
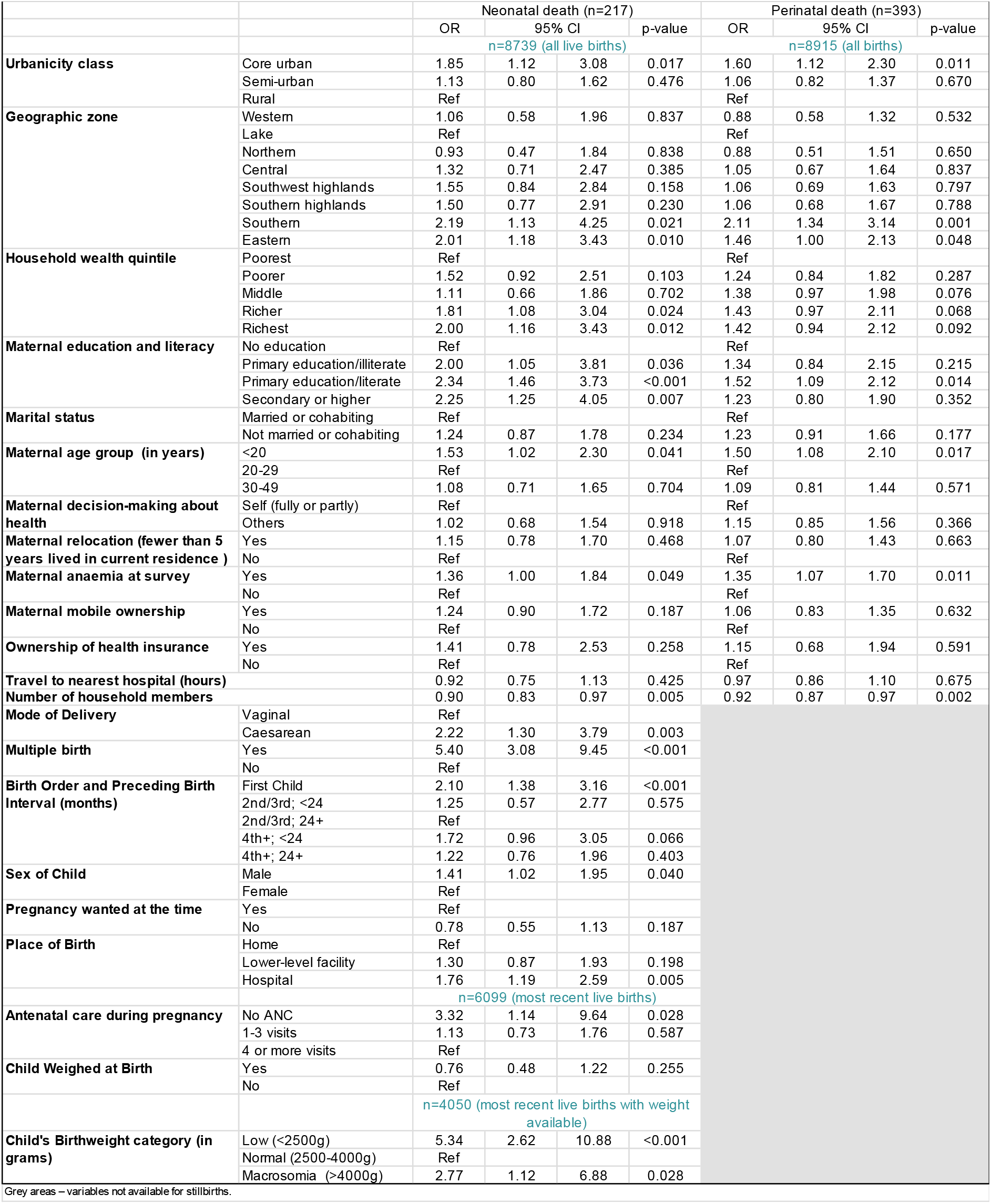
Bivariate associations with neonatal death and perinatal death.

### Multivariable analysis

Table 5 shows the results of the four multivariable models. Overall, these models show that the adjusted odds of neonatal death in core urban areas was between 26% and 136% higher, and in semi-urban areas 26%-77% higher compared to rural areas. The adjusted odds of perinatal death in core urban areas were 71% higher and in semi-urban areas 8% higher compared to rural areas. The direction of association was consistent across the four models, but in none of them was it significant at the <0.05 level. The main objective of this paper was to assess the strength of the association between urbanicity categories and neonatal/perinatal mortality after adjustment for confounding by other factors. However, we briefly report important findings about the association between other variables in the multivariable models and odds of death. Variables consistently and strongly associated with higher odds of neonatal and/or perinatal death included residence in the Southern zone, maternal anaemia at time of survey, smaller household size, multiple birth, primiparity and short birth interval, male sex of the child, and both low and high birth weights. Additional variables associated with higher adjusted odds of neonatal or perinatal death, but with inconsistent levels of significance across the models, were: higher maternal education, younger maternal age, and non-use of antenatal care. Travel time to the nearest public hospital, household wealth quintile, and type of facility where birth took place were included in the models as confounders, but were not associated with neonatal or perinatal mortality.

**Table 5.**
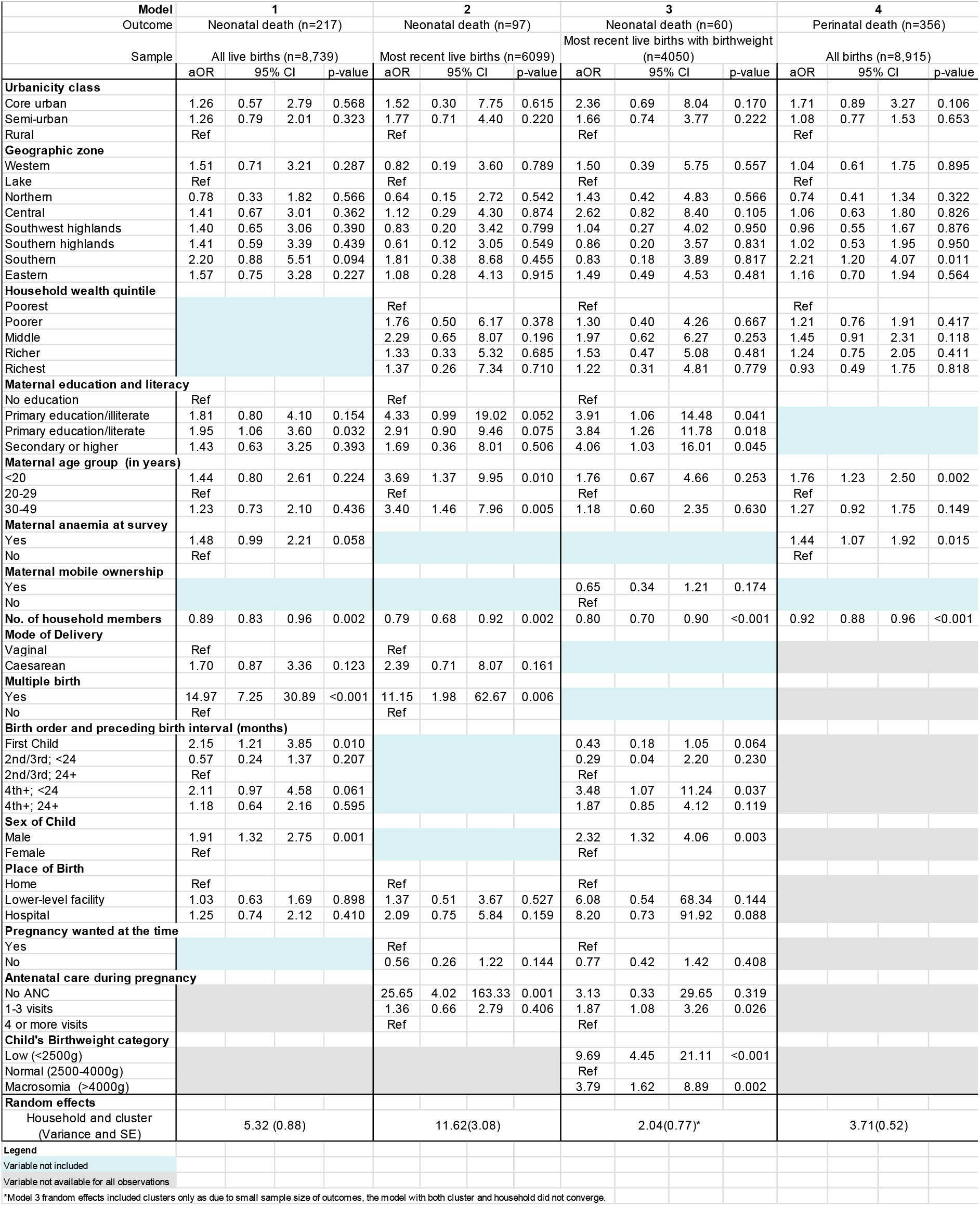
Multivariable associations with neonatal death and perinatal death (four models)

## Discussion

Our analysis based on DHS data from mainland Tanzania in 2015-16 showed that the three GHS-SMOD urbanicity categories captured the meaning of urbanicity more accurately than the DHS urban/rural residence, because they were derived from satellite data and based on built environment and population density, rather than on administrative delineation and unclear cut-offs. The most important cause of misclassification between the two methods was that some clusters considered urban by DHS were rural according to GHS-SMOD. Further, we disaggregated to core urban and semi-urban allowing more accurate capture of the variation in human settlement on a continuum and to expose any dose-response associations. The new category of semi-urban had levels of neonatal and perinatal mortality similar to rural areas. Next, we assessed whether the urbanicity categories were associated with both neonatal and perinatal mortality, thus addressing some misclassification between neonatal deaths and stillbirths. We carefully adjusted for confounding using variables collected by the DHS and an additional variable capturing travel time to nearest public hospital. Further, we used random effects to account for the clustering on household and area levels. We ran four separate models on sub-samples with varying availability of confounders. The results of these analyses showed a consistent pattern of higher odds of neonatal and perinatal death with increasing levels of urbanisation and similar effect size reported by Norris et al.^7^ However, the associations were not significant at the p<0.05 level, most likely due to a small sample size of neonatal and perinatal deaths. Taken together, these findings bolster our confidence in the evidence showing an association between higher levels of urbanicity and higher neonatal and perinatal mortality. In the Tanzanian context, this effect was driven predominantly by core urban areas. We discuss several findings from our study to expound potential mechanisms underlying this association.

A recent paper on the reasons for loss of urban mortality advantage among adults (15-49 year olds) from multiple DHS surveys using the urban/rural stratification noted that rapid expansion of population in slums has led to premature mortality linked to overcrowding, poverty, road traffic accidents, lack of sanitation and the double burden of malnutrition leading to non-communicable diseases in this population.^4^ On the other hand, they described an urban advantage in child survival, which they attributed in part to better access to healthcare, better infrastructure, greater economic opportunities and other factors such as lower fertility levels and longer birth intervals. Causes of stillbirths and deaths in the neonatal period are a combination of the various factors affecting the health of adults in general and pregnant women in particular (e.g., maternal nutrition, exposure to infections such as STIs and malaria, occupational hazards, exposure to heat and pollution), as well as that of children (access to healthcare and the quality of that care, particularly at time of labour and birth).

Multiple causal pathways for the effect of urban residence on neonatal survival have been proposed, including individual health-seeking behaviour/accessibility of care, obstetric risk factors, quality of care during pregnancy, childbirth and the postnatal period, as well as broader issues related to socioeconomic determinants, urban living conditions and urbanisation processes.^7^ Our multivariable models included variables capturing all these four dimensions. While some of these were significantly associated with neonatal and perinatal mortality, their inclusion did not completely explain the association between urbanicity and neonatal/perinatal mortality. We highlight several findings which could inform future analyses to explore the causal pathways in more depth.

Issues linked to access to care and care quality in urban areas are numerous. The use of ANC and facility-based childbirth care in large cities in Africa is near-universal (>94% in Dar es Salaam), but characterised by high levels of private sector use and inconsistent receipt of evidence-based interventions.^42^ The analysis of 22 large African cities also showed variable levels of essential care elements (>99% of babies born in health facilities were weighed but only half of babies initiated breastfeeding within an hour of birth) and high levels of early discharge from health facilities following both vaginal and caesarean section births in Dar es Salaam. Further, literature shows that poor women, especially those living in informal settlements, might also receive poorer quality of care, encounter stigmatising attitudes and disrespectful care in health facilities.^43-45^ Our additional analysis on the timing of deaths showed that a comparatively low percentage of neonatal deaths in core urban areas occurred on the day of birth compared to semi-urban and rural areas. Interpretation is difficult, but one explanation may be better access to emergency obstetric care including neonatal resuscitation in core urban compared to rural areas.^46^ Some resuscitated babies may still die a few days later because of underlying conditions due to complications of preterm birth, infections, and late complications from asphyxia.

Low birthweight was associated with a nearly 10-fold increase in neonatal mortality. The high degree of underreporting of stillbirths in the core urban area points to potential misclassification of stillbirths as neonatal deaths or general underreporting of stillbirths in these contexts. Misclassification in household surveys has been reported in several studies^13 47-49^ and is related to factors such as how well vital status is assessed at birth and birth attendant’s ability to distinguish stillbirths from early neonatal deaths, socio-cultural perceptions of pregnancy loss that might affect disclosure, or intentional misclassification or underreporting to avoid blame or additional investigation depending on requirements in the context. That this pattern appears confined only to urban areas in our study warrants further investigation. We would expect better differentiation between these outcomes in an urban setting where higher quality services and more skilled personnel are available. Another contributing factor could be the impact of recent training on neonatal resuscitation in several health facilities in these areas which may be improving the survival of babies thus leading to fewer stillbirths.

On the other hand, modelled travel time to nearest hospital was neither associated with neonatal nor perinatal mortality. In the sample of live births, one third of women reported giving birth in hospitals, but this is a combination of women who planned to give birth there and those referred to hospitals due to a maternal or fetal complication. Rather than hospitals being a cause of higher neonatal mortality,^50^ the more complicated case mix in hospitals might be reflected in the higher adjusted odds of neonatal death in this facility type (which was not significant). Previous studies show that facility readiness is comparable across urban and rural settings in Tanzania.^51^ Birth by caesarean section was associated with 70%-139% higher odds of neonatal death, but not significant which is most likely due to small sample size of births by caesarean section. It is likely that the neonatal death is a result of a complication, and birth by caesarean section is a clinical intervention in response to the same complication, meaning that caesarean section is a proxy for complications rather than the proximal cause of neonatal death.^52^ Next, while only 2% of women reported not receiving any ANC, this category had higher odds of neonatal mortality compared to women receiving four or more ANC visits. While non-use of ANC was rare and might be a marker of general socio-economic disadvantage, it also means that women did not receive essential care elements linked to reduced maternal and perinatal mortality and morbidity, including malaria screening and preventive therapy as well as screening and treatment for conditions linked to stillbirths and prematurity such as sexually transmitted infections, eclampsia and anaemia.

Two socio-economic variables were associated with the outcomes – maternal education and household size. The three models looking at neonatal mortality showed a consistent association between higher levels of education and *higher* neonatal mortality. This is unlikely to be a result of confounding by older maternal age (which is linked to poorer perinatal survival^53^) because our multivariable model includes this variable. One possible explanation is that the extent of under-reporting of neonatal deaths is higher among women with no education because of stigma^54^, thus artificially increasing the odds of mortality among those with higher levels of education. Some of these deaths might be misclassified as stillbirths, resulting in the finding that maternal education was not an important confounder in the perinatal mortality model and was excluded. Higher number of household members was consistently and significantly associated with lower adjusted odds of neonatal and perinatal mortality. We estimate that for every additional household member, the odds of neonatal and perinatal mortality declined by approximately 10%. This points to the importance of familial support including advocating and enabling timely care-seeking (e.g., by recognising danger signs, providing childcare during woman’s absence, or assisting during travel), help within the household, and with enabling positive behaviours such as self-care and breastfeeding.^55^ We did not explore the type of household members which were essential to this decrease in mortality. It is possible that the presence and role of a male partner is different to the roles played by trusted and experienced females such as mothers, sisters, mothers-in-law and aunts. Their presence, availability and support is likely to differ by area type.

We identified several known biological risk factors which are linked to increased neonatal and perinatal mortality in the absence of accessible, high-quality care. All four models showed that being <20 years and being 30 years old or older, compared to being 20-29 at time of birth, was associated with higher adjusted odds of neonatal/perinatal mortality (in the model which includes stillbirths, this association was highly significant). Second, maternal anaemia, even though measured at the time of survey which could be several years after the births, was associated with an approximately 50% increase in both neonatal and perinatal mortality. High prevalence of anaemia in Tanzania is shown and our analysis confirms the link to pregnancy/childbirth complications.^56^ Third, male newborns had approximately double the odds of neonatal death compared to female newborns, again, a well-known risk factor reflecting biological vulnerability of males.^57^ However, the size of the effect also suggests that there might be under-reporting of female neonatal deaths, which had been documented in other settings with cultural preferences for male children. Fourth, multiplicity increased the odds of neonatal mortality 11-15-fold, in line with the known evidence.^58^ This is a known risk factor which can be mitigated by timely diagnosis of multiple pregnancy and continuous care during pregnancy, childbirth and the early postnatal period. Fourth, we also note that first birth and birth after a short birth interval at high parity (4 or more) were associated with higher neonatal mortality. It is possible that the manner in which these known and yet unknown risk factors operate is different in densely populated urban settings compared to rural areas. While the sample size available on the DHS did not allow us to test for interactions, we note that improving access to good quality care is essential for preventing neonatal and perinatal deaths.

### Strengths and limitations

Our in-depth analysis of the association between urban residence and neonatal and perinatal mortality addressed several critical limitations of previous studies. We were able to more accurately classify the extent of urbanicity.^59^ However, our indicator of urbanicity has limitations, including grouping affluent parts together with slums or informal settlements in core urban. The alternative could have been to use a composite measure combining wealth quintile and urbanicity to construct a fourth category referring to slums and informal settlements – proxied by the poorest quintiles living in core urban areas slums.^60^ However, sample size constraints of the main outcomes made this approach unfeasible. By including the two outcomes of neonatal mortality and perinatal mortality, we addressed some effects of the proposed misclassification between stillbirth and neonatal deaths. Still, our data indicate that neonatal and perinatal deaths are underreported in these survey self-reports, in view of the implausible higher neonatal mortality in better educated and more wealthy groups as well as the much higher odds in male births. Limitations also exist in several other variables. Travel time was based on the nearest hospital, whereas in reality, women often bypass the nearest facility.^61 62^ Further, we made assumptions about travel speed, which may not hold true in all places and might have a larger margin of error within cities due to, for example, variability in traffic and weather, and waiting time.^63^ However, this was necessary due to lack of observational data.^64^. The exact location of the household of residence for each woman is obscured by provision of one cluster location and by cluster displacement in DHS due to reasons of anonymity. We tried to ameliorate this by including some cluster level variables which would tell us about the lived environment of the ‘neighbourhood’.

We were unable to include an indicator for malaria as the DHS does not include a specific assessment in pregnancy. Malaria may be one factor to explain the regional differences, for example the higher mortality in the Southern region compared to high altitude regions of Northern and Southwestern and Sothern Highlands. It was not possible to perform a more in-depth analysis for stillbirths as important explanatory variables are not collected and therefore, not accounted for in the model. Also, other key variables were missing depending on the outcome chosen (see table 5) meaning that none of the four models were theoretically complete. Our Model 3 does not have fixed effects for household instead additional key variables (Birth weight and Number of ANC visits) captured some differences between the households that were being captured by the random effect. The data excludes the sample of babies of women who died themselves, as these neonatal and perinatal deaths were not captured in household surveys. Finally, even though the DHS is a nationally representative survey and the number of women interviewed had increased in recent years, the sample size of neonatal deaths and stillbirths was relatively small. The limited sample size could be one reason why we did not detect a significant association between urbanicity and mortality in the multivariable results.

## Conclusion

In our advanced analysis which improved the accuracy of the exposure variable (urbanicity), reduced reporting bias in outcome (by adding stillbirths) and adjusted for confounding and clustering more completely, we found moderate evidence of higher neonatal and perinatal mortality in semi-urban and particularly in core urban areas compared to rural areas in mainland Tanzania. The effect seemed to follow a dose-response pattern with increasing extent of urbanicity. This is consistent with earlier findings, and might extend to other countries with slower neonatal mortality declines in urban areas as noted by Norris et al. Our multivariate analysis aimed to provide an in-depth understanding of the mechanisms of this association, however, we appreciate that many questions are still unanswered due to the data limitations. Therefore, we call for more in-depth analyses to disentangle the contribution of pregnancy factors, living conditions and quality of care in birthing facilities.

Addressing the high rates of mortality in urban areas is also critical for Tanzania to meet the SDG target on reduction of NMR to less than 16/1,000 live births by 2030. Focusing solely or predominantly on rural areas is unlikely to tackle the high and largely preventable neonatal and perinatal mortality identified in urban areas, whether in the core, densely populated urban centers and particularly informal settlements, or the growing semi-urban areas around Tanzania’s main and secondary cities and towns. In order to appropriately target interventions, we must rely on more up-to-date, accurate and granular capture of urbanicity, which is possible through using innovative satellite technologies and spatial epidemiology approaches. We call for better data allowing disaggregation’s into neighbourhoods of slums and informal settlements to ascertain whether across communities the ‘urban’ category is masking heterogeneities.

## Data Availability

DHS datasets are available from the DHS programme upon request at https://dhsprogram.com/data/available-datasets.cfm. The health facility database used to compute travel time in Tanzania is available at https://doi.org/10.6084/m9.figshare.c.4399445 while the urbanicity surfaces can be accessed at http://data.europa.eu/89h/42e8be89-54ff-464e-be7b-bf9e64da5218

https://doi.org/10.6084/m9.figshare.c.4399445

http://data.europa.eu/89h/42e8be89-54ff-464e-be7b-bf9e64da5218

## Author contributions

Conceptualization: PMM, LB, CH, JP; Investigation: PMM, LB, JP, AS, AC, ABP, CH; Data curation: PMM, JP, LB; Formal analysis: PMM, LB; Visualisation: PMM, LB, AS; Writing – Original Draft Preparation: PMM, LB, JP, AS, AC, CH; Writing – Review & Editing: PMM, LB, JP, AS, AC, ABP, CH.

## Acknowledgments

We would like to thank the DHS team, the survey enumerators and the women who contributed information about their lives. We also acknowledge discussions with Dr Ulrika Baker (UNICEF Tanzania) and with Cameron Taylor on DHS measurement of urban/rural residence.

