## Supplementary file 1 for "Neonatal and perinatal mortality in the urban continuum: A geospatial analysis of the household survey, satellite imagery and travel time data in Tanzania"

Table 1: The mean travel time to the nearest hospital from each cluster stratified based on DHS (urban and rural) and GHS-SMOD (core urban, semi-urban and rural) classifications in Tanzania. The DHS and GHS-SMOD represents the same spatial extents

| Source | Urban/rural classification | Number of clusters | Mean travel time (minutes) |
| --- | --- | --- | --- |
| **DHS** | Urban | 163 | 14.4 |
|  | Rural | 364 | 78.2 |
| **GHS-SMOD** | Core urban | 61 | 4.3 |
|  | Semi-urban | 224 | 41.1 |
|  | Rural | 242 | 89.4 |
| **National** | 8915 | 527 | 62.8 |

Figure 1**.** The spatial distribution of the seven classes based on the 2015 GHS-SMOD layer in Tanzania describing the continuum from urban to rural areas in 2015.

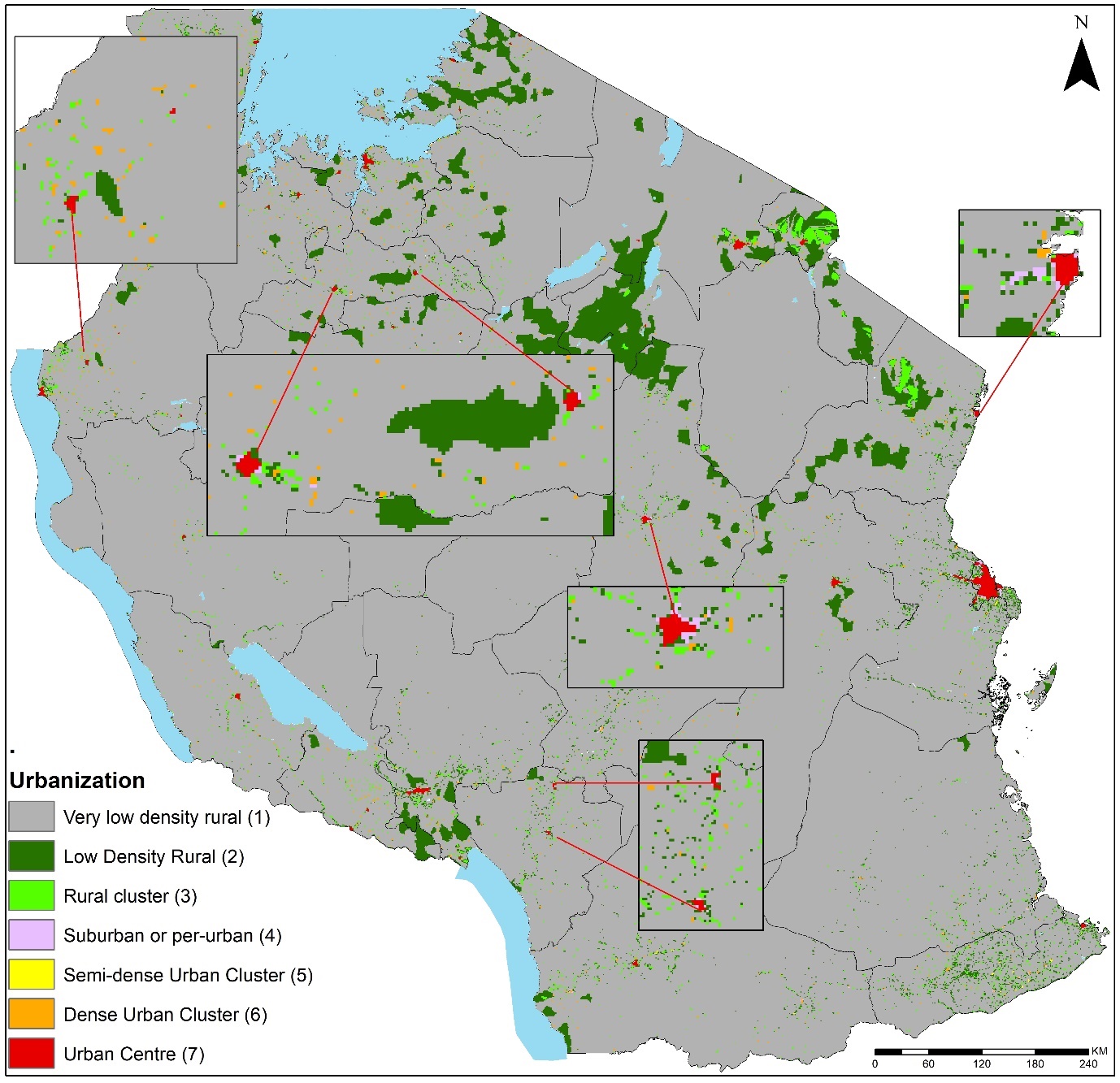

Figure 2. GHS-SMOD-derived urbanicity classifications based on satellite imagery

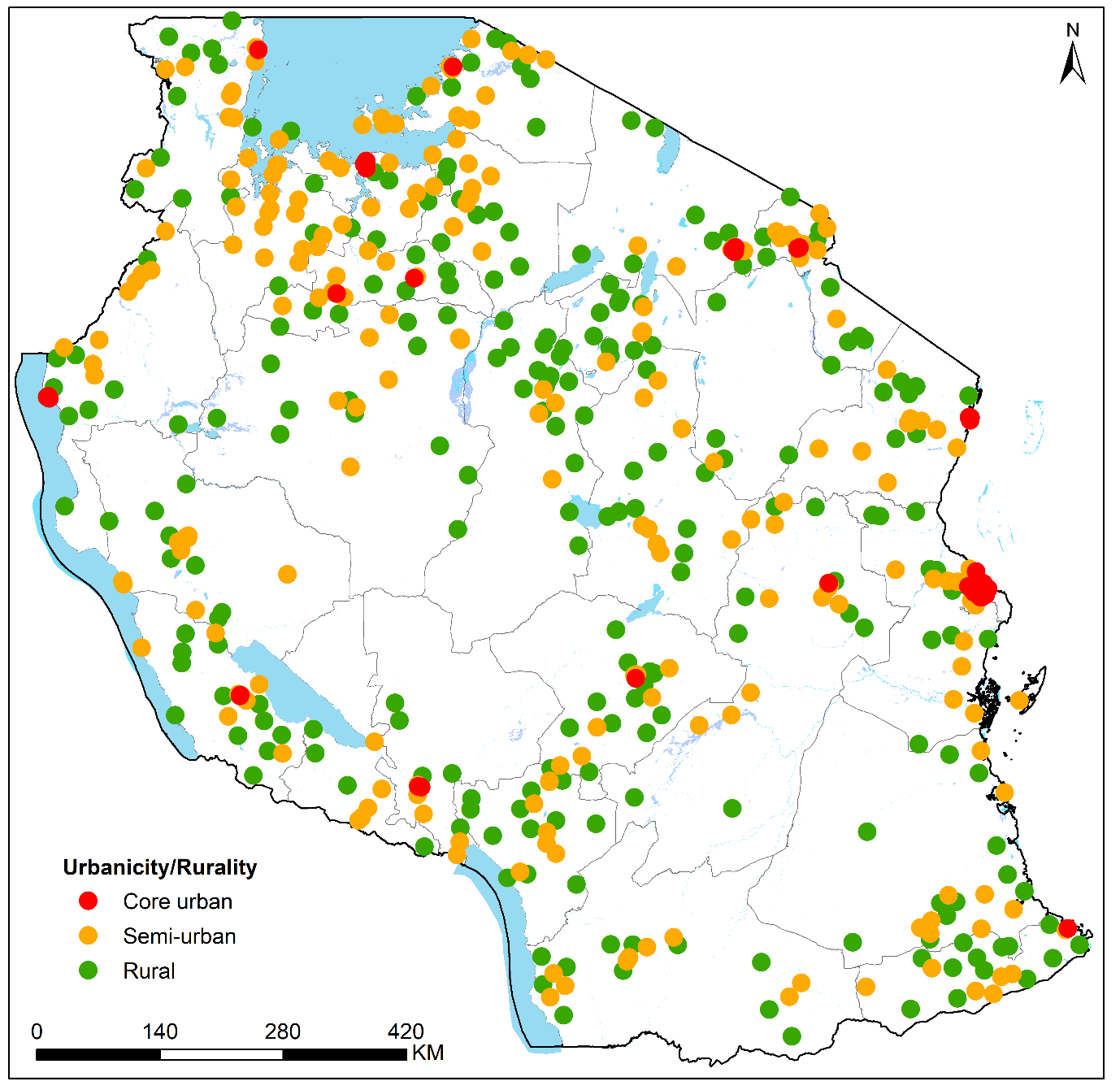

Figure 3. Travel time to the nearest hospital (n=236) in Tanzania in 2015 at 300m spatial resolution classified into 5 classes ranging from less than 30 minutes (green) to over 2 hours/120 minutes (red). The white areas are national parks/reserves that were considered as barriers except in presence of roads.

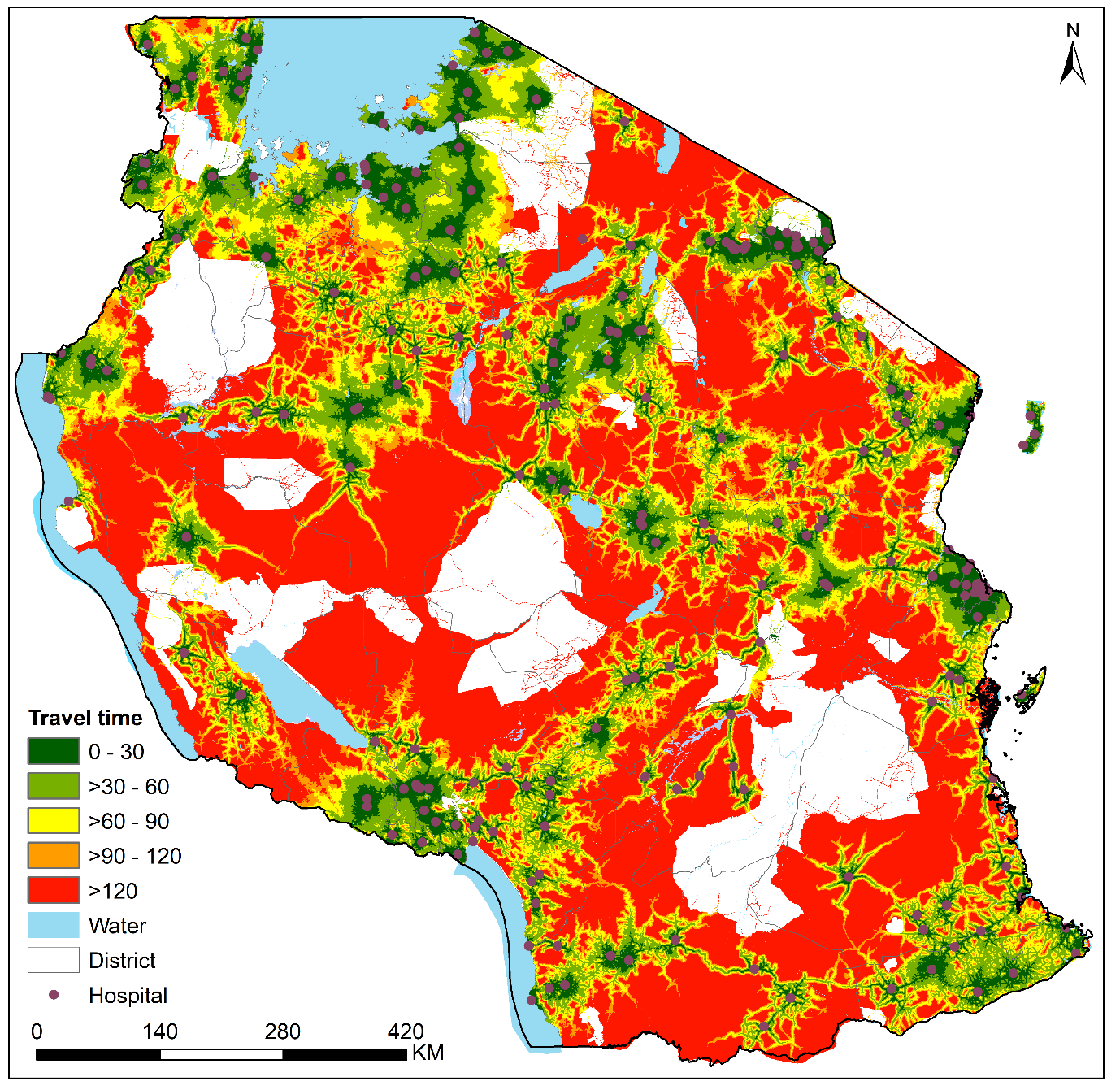

Figure 4. Travel time to the nearest hospital (n=236) in Tanzania in 2015 at cluster level classified into five classes ranging from less than 30 minutes (green) to +120 minutes (red)

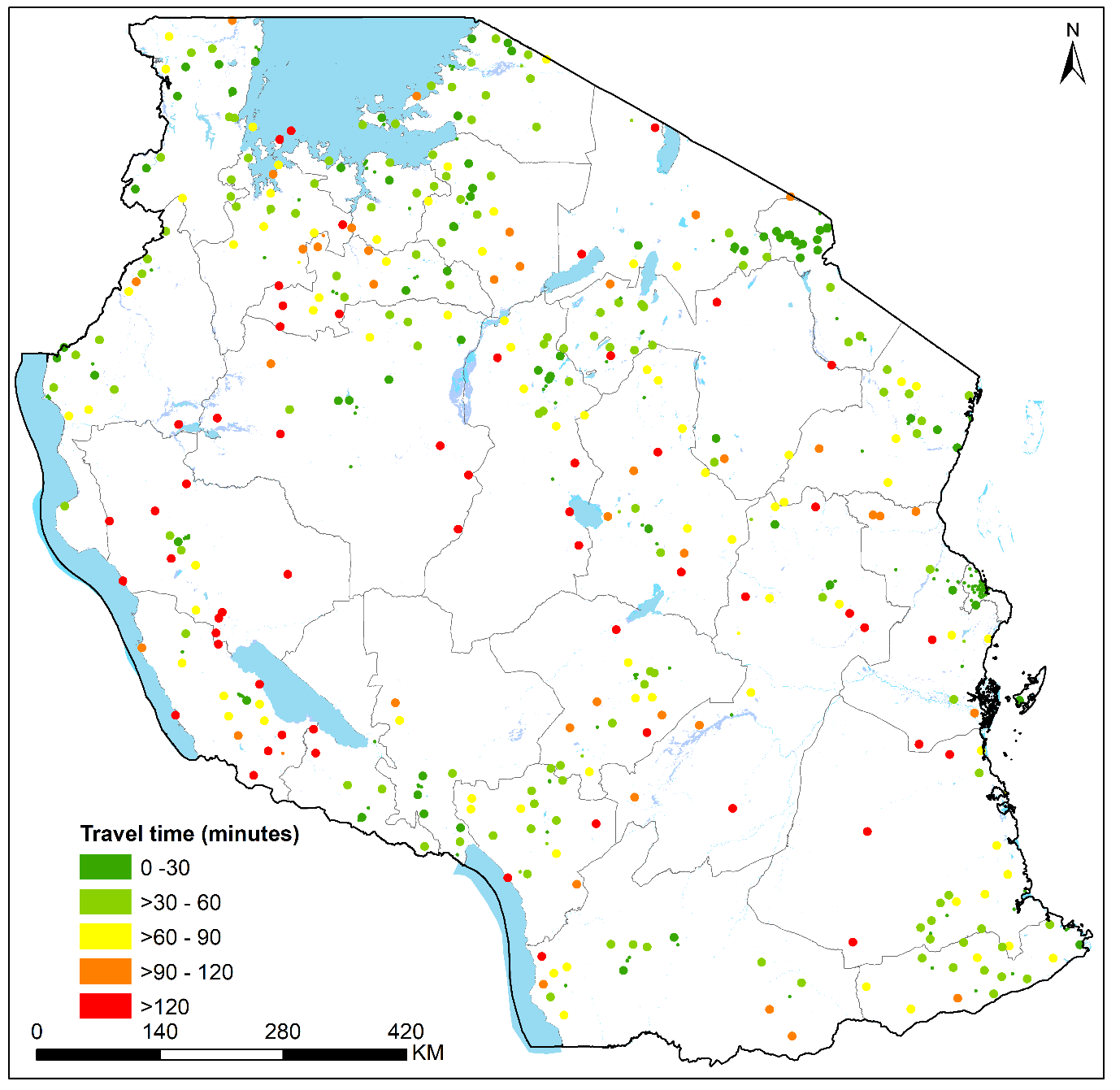
