## Supplementary figures and images for "Neonatal and perinatal mortality in the urban continuum: A geospatial analysis of the household survey, satellite imagery and travel time data in Tanzania"

### Supplementary file 2

#### Supplementary file 2

**Summary of variables used in analysis**

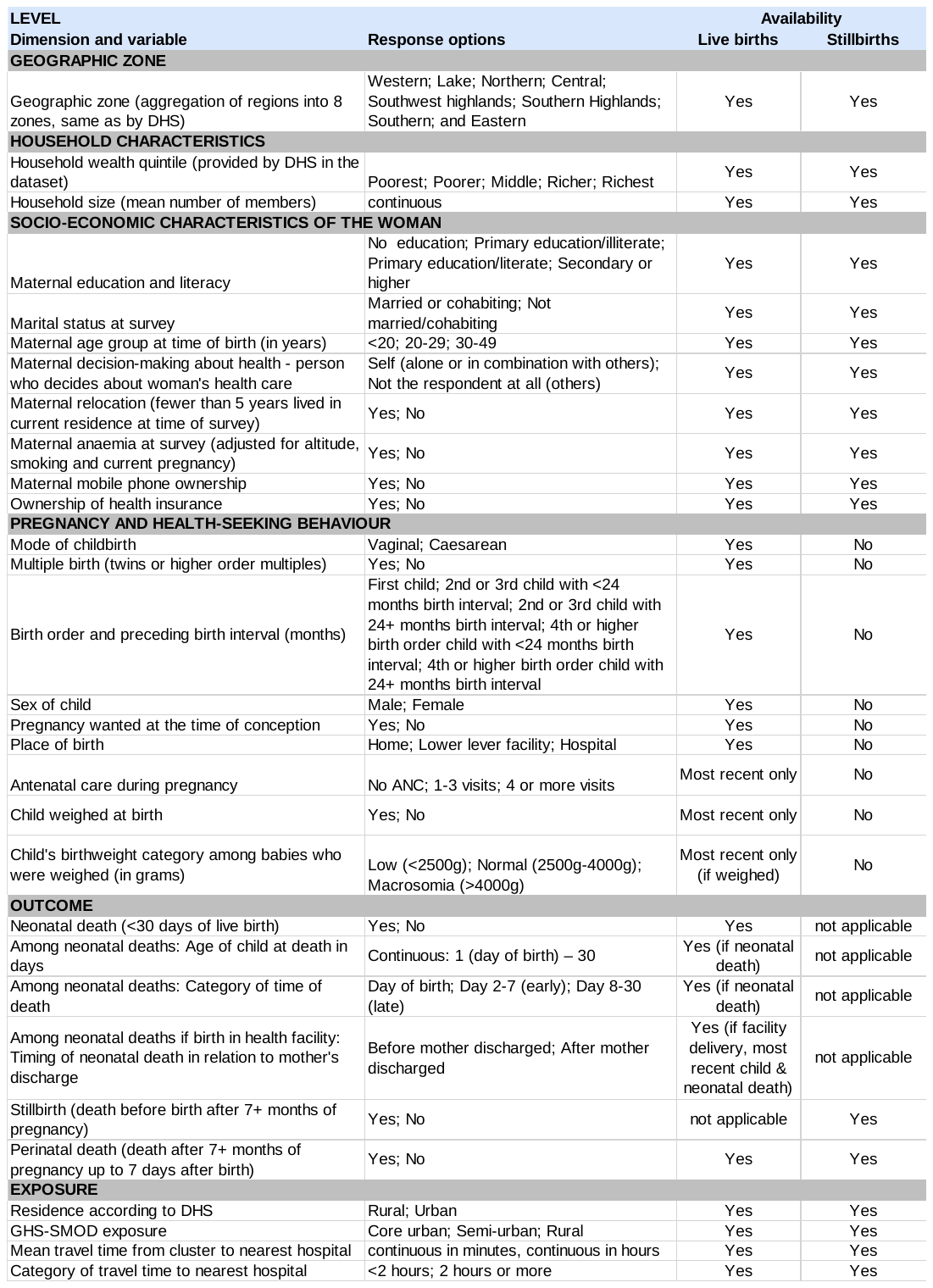
