## Supplementary file 3 for "Neonatal and perinatal mortality in the urban continuum: A geospatial analysis of the household survey, satellite imagery and travel time data in Tanzania"

**Distribution neonatal deaths by age at death and facility discharge, by urbanicity category**

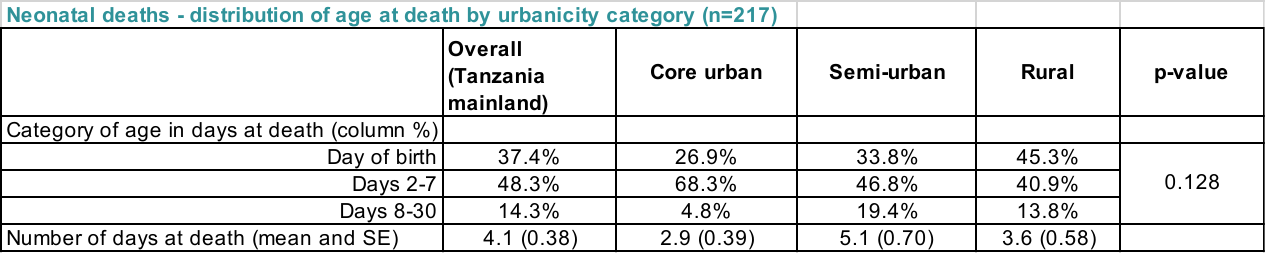

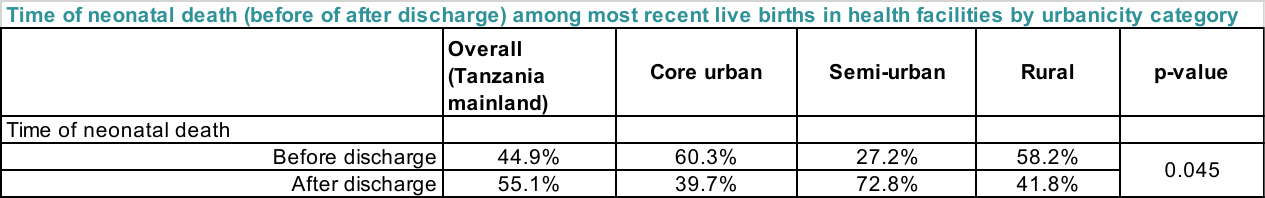

**Stillbirths: Early neonatal death ratio**

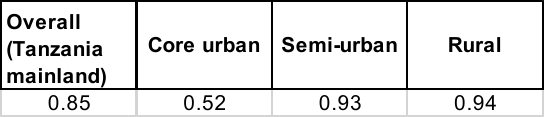
